# Estimation of novel coronavirus (covid-19) reproduction number and case fatality rate: a systematic review and meta-analysis

**DOI:** 10.1101/2020.09.30.20204644

**Authors:** Tanvir Ahammed, Aniqua Anjum, Mohammad Meshbahur Rahman, Najmul Haider, Richard Kock, Md. Jamal Uddin

## Abstract

Understanding the transmission dynamics and the severity of the novel coronavirus disease 2019 (COVID-19) informs public health interventions, surveillance, and planning. Two important parameters, the basic reproduction number (*R*_*0*_) and case fatality rate (CFR) of COVID-19, help in this understanding process. The objective of this study was to estimate the *R*_*0*_ and CFR of COVID-19 and assess whether the parameters vary in different regions of the world. We carried out a systematic review to retrieve the published estimates of the *R*_*0*_ and the CFR in articles from international databases between 1^st^ January and 31^st^ August 2020. Random-effect models and Forest plots were implemented to evaluate the mean effect size of the *R*_*0*_ and the CFR. Furthermore, the *R*_*0*_ and CFR of the studies were quantified based on geographic location, the tests/thousand population, and the median population age of the countries where studies were conducted. The I^2^ statistic and the Cochran’s Q test were applied to assess statistical heterogeneity among the selected studies. Forty-five studies involving *R*_*0*_ and thirty-four studies involving CFR were included. The pooled estimation of the *R*_*0*_ was 2.69 (95% CI: 2.40, 2.98), and that of the CFR was 2.67 (2.25, 3.13). The CFR in different regions of the world varied significantly, from 2.51 (2.12, 2.95) in Asia to 7.11 (6.38, 7.91) in Africa. We observed higher mean CFR values for the countries with lower tests (3.15 vs. 2.16) and greater median population age (3.13 vs. 2.27). However, the *R*_*0*_ did not vary significantly in different regions of the world. An *R*_*0*_ of 2.69 and CFR of 2.67 indicate the severity of the COVID-19. Although *R*_*0*_ and CFR may vary over time, space, and demographics, we recommend considering these figures in control and prevention measures.

## Introduction

Since the emergence of SARS-CoV-2, the virus causing COVID-19, in late 2019, there have been more than 35 million confirmed cases, and 950,000 confirmed deaths have been reported as of 19 September 2020 [1]. The basic reproduction number (*R*_*0*_), used to measure the transmission potential of a disease, can be used to inform public health policies. The *R*_*0*_ is defined as the expected number of cases directly generated by one infected case in a population where all individuals are susceptible [2,3]. Outbreaks will continue to propagate until this number falls below 1.0. A timely and accurate assessment of COVID-19’s *R*_*0*_ informs COVID is of great significance for assessing the spread of the virus, predicting future trends, and adjusting a series of control measures.

Another important factor to inform is the case fatality rate (CFR), which is the proportion of deaths compared to the total number of people diagnosed with a disease within a defined population of interest and is typically presented as a percentage [4]. CFR is an indicator of disease severity and can be used to identify high-risk populations to inform targeted interventions and the overall public health response [5].

To understand the transmissibility and severity of SARS-CoV-2 in populations, there have been several estimates of *R*_*0*_ and the CFR. Estimates of R_0_ and CFR have varied in different countries and different settings [6]. For example, in the Diamond Princess cruise ship, the estimated median *R*_*0*_ was 2.28 (95% CI: 2.06-2.52) [7]. In Wuhan, during the early phase of the outbreak, the *R*_*0*_ was estimated as 2.24 (95% CI: 1.96-2.55) [8]. The *R*_*0*_ has also been estimated for several other countries including Italy, 3.2 (95% CI: 2.9-3.5) [9]; USA, 5.3 (95% CI: 4.35-6.25) [10]; and, Japan, 2.6 (95% CI: 2.4-2.8) [11].

Likewise, estimates of CFRs have varied in different countries and phases of the pandemic. A study found that at the start of the epidemic, CFR in China was 2%, but on 5 March 2020, it was increased to 3.7%. In South Korea, the same study reported the CFR as 0.6% on 5 March 2020 [12]. In Europe, during March 2020, Italy, Spain, and France had a CFR of 9.26%, 6.16%, and 4.21%, respectively [13]. Given the considerable variation in estimates of both these parameters, in this current study, our aim was to conduct a systematic review and meta-analysis of the published literature on the COVID-19 outbreak to provide summary statistics of the *R*_*0*_ and the CFR, which have the best applicability for other countries or regions.

## Methods

We followed the Preferred Reporting Items for Systematic Review and Meta-Analysis Protocols (PRISMA-P) 2009. We registered the study protocol with the International prospective register of systematic reviews (PROSPERO) (registration number ID: CRD42020182867).

### Search strategy

We systematically searched the major databases: LitCovid (a curated COVID-19 database of PubMed), PubMed, EMBASE, Scopus, and Web of Science databases from 1 January 2020 to 31 August 2020. We searched articles for “Basic reproduction number (*R*_*0*_)” and “Case-fatality rate (CFR)” of the COVID-19 patients. For basic reproduction number, the search terms were: “2019 novel coronavirus and COVID-19” AND “Basic reproduction number”, OR “Basic reproduction rate” OR “*R*_*0*_”. For the case fatality rate, the search terms were “2019 novel coronavirus and COVID-19” AND “case fatality rate”, OR “case fatality ratio”, OR “CFR” according to the indices of the various databases (Supplementary Table S1). We also hand searched included papers’ reference lists and contacted experts in the field to ensure a comprehensive review. Records were managed by Mendeley version 1.19.4 software to exclude duplicates.

### Inclusion and exclusion criteria

Articles were selected if they reported CFR and/or R0 with a confidence interval of the patients with COVID-19. The inclusion criteria for studies were laboratory-confirmed parents with COVID-19; reported at sufficient sample size; considered all age-groups; and published in English and any counties or regions of the world. Studies were excluded if they were: pre-prints; reviews articles; editorials and case reports; letters to the editor and short communications; and specific to children or pregnant women. However, we did not exclude some special populations representative of the general population in a particular setting (e.g., Diamond Princess cruise ship). The steps of the literature search are illustrated in Fig. 1.

**Fig. 1.**
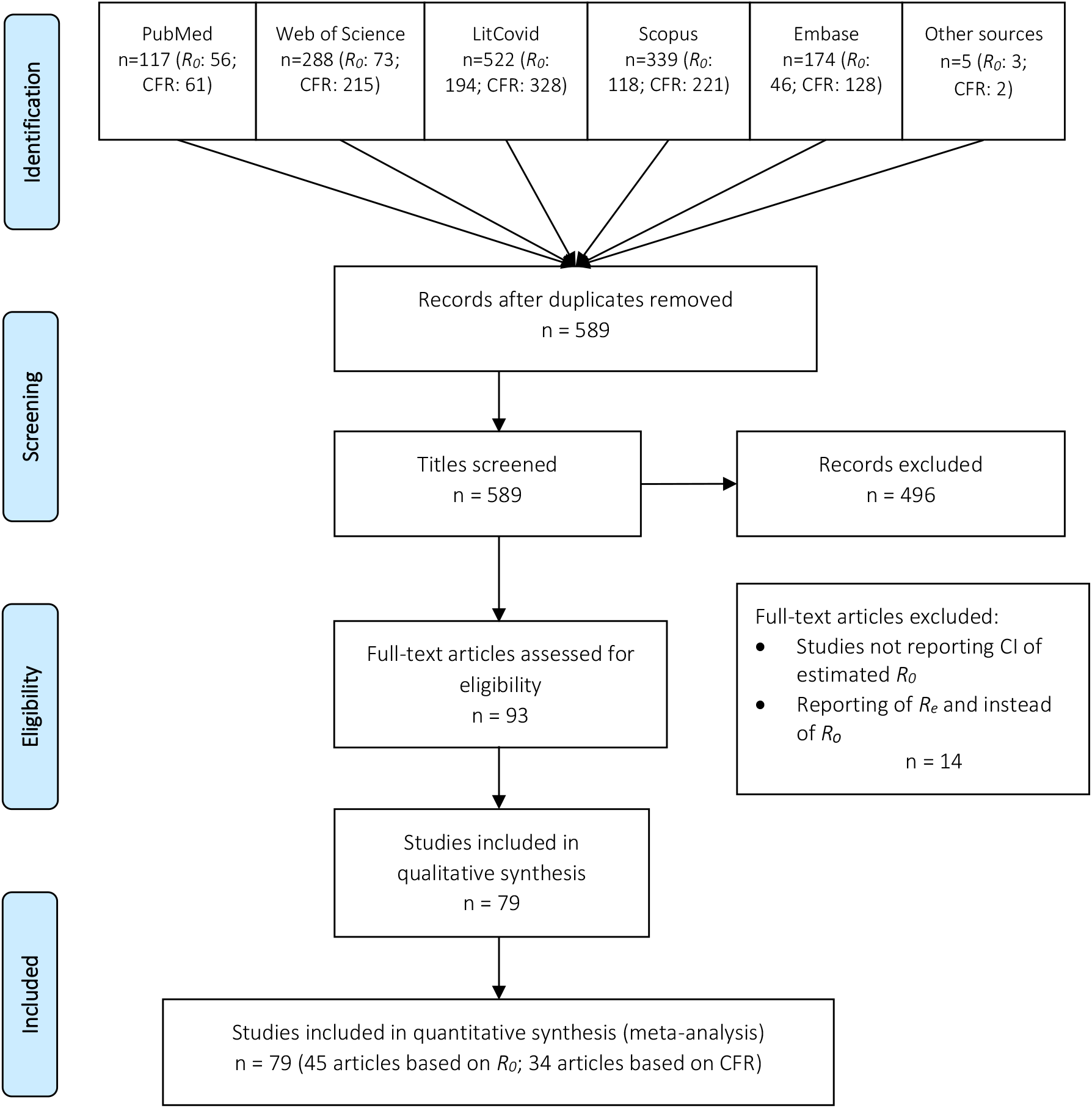
Flow diagram for included studies in the meta-analysis. CFR = Case Fatality Rate; *R*_*0*_ = basic reproduction number; *R*_*e*_ = effective reproduction number.

### Data screening and extraction

After excluding duplicate papers, two investigators (TA and AA) independently screened the titles and abstracts using the eligibility criteria and then assessed the rest full-text articles for eligibility. Disagreements were resolved through discussion with co-authors. A standardized form was used to extract data from all eligible studies. For each selected articles, the following information’s were extracted: publication details [title, first author, publication year, journal name and publisher]; design and population [country, study design, study period, and sample size]; participants’ characteristics and major findings [number of deaths, case fatality rate, basic reproduction number, confidence interval, methods applied, standard deviation, and any other relevant statistics]. Forty five research articles [7,9,20–29,10,30–39,11,40–49,14,50–54,15–19] for *R*_*0*_ and 34 research articles [13,15,62–71,24,72–81,55,82–85,56–61] for CFR estimation were selected.

### Critical appraisal and publication bias

The critical appraisal of each selected study was assessed by TA and AA independently using modified Risk of Bias (ROB) operational criteria (Supplementary Table S2 and Table S3) [86]. Publication bias was assessed using a funnel plot, and the Egger’s test with P < 0.05 was an indication of small-study effects.

### Statistical analysis

We defined *R*_*0*_ as the expected number of cases directly generated by one infected case in a population where all individuals are susceptible and CFR as the percentage of COVID-19 cases that result in death. Background statistics of the COVID-19 patients were recorded through frequency distribution analysis. A random-effect model was, therefore, used to perform a meta-analysis. The geo-location variations for *R*_*0*_ and geo-location, tests per thousand population, and population median age variations for CRF were examined using subgroup analysis. The median values of the frequency distributions were used as cut-off values of the sub-groups. Heterogeneity was assessed among the selected studies using the Higgin’s & Thompson’s I^2^ statistic and Cochran’s Q test. All statistical analyses were performed using Stata SE version 16.1 (Stata Corporation, College Station, TX, USA 5.

## Results

We identified a total of 490 articles estimating *R*_*0*_. After considering the duplicates, inclusion, and exclusion criteria, we reviewed these research articles. We searched for titles, abstracts and keywords, methodology, and references and found 59 articles to investigate further (Fig. 1). Out of these 59 articles, we found 45 research articles that reported 108 *R*_*0*_ estimates ranging from 0.27 to 8.21 with related statistics for a meta-analysis (Supplementary Table S4). Similarly, for the CFR, after considering filtering criteria, only 34 research articles out of 955 articles qualified for quantifying parameter values of CFR (Fig. 1), and these 34 articles provided 158 estimates of CFR ranging from 0.00 to 25.9 for use in the meta-analysis (Supplementary Table S5). No studies were excluded.

### Pooled estimation of the basic reproduction number (*R*_*0*_)

In the random-effects model, the pooled *R*_*0*_ for COVID-19 was estimated as 2.69 (2.40, 2.98), indicating that an infected person with COVID-19 can transmit the infection to on average three susceptible people (Table 1, and Fig. 2). There was significant heterogeneity between studies (I^2^: 99.96) (Table 1).

**Table 1.**
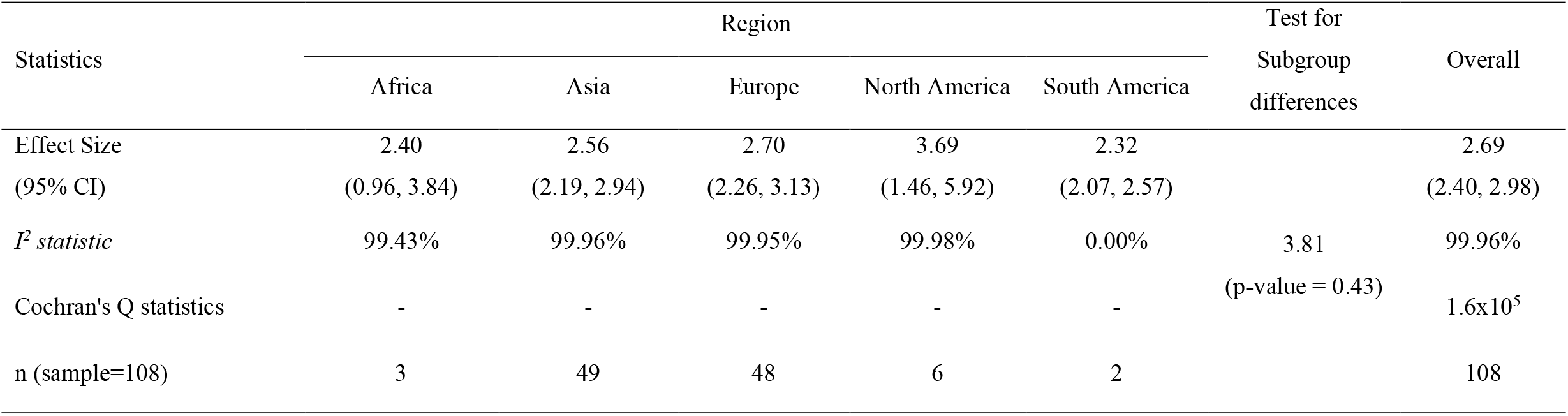
Effect size and test of heterogeneity for the basic reproduction number (*R*_*0*_) by region.

**Fig. 2.**
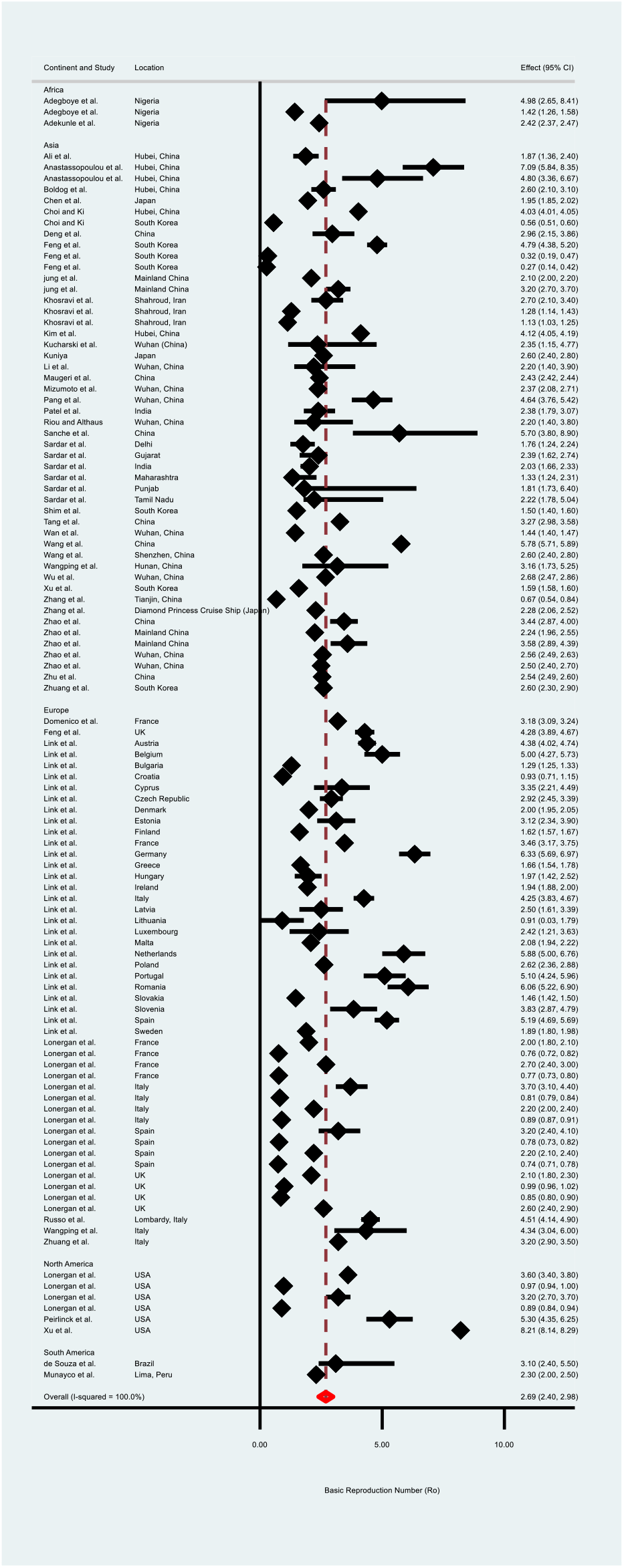
Forest plot of the basic reproduction number (*R*_*0*_) values based on the random-effects model.

The outcome of subgroup analysis showed that the *R*_*0*_ value for Africa was 2.40 (0.96, 3.84), for Asia was 2.56 (2.19, 2.94), for ‘Europe’ was 2.70 (2.26, 3.13), for North America was 3.69 (1.46, 5.92), and for South America was 2.32 (2.07, 2.57) (Table 1). However, the test of regional subgroup differences using the random-effects model confirms that the estimated mean *R*_*0*_ values are not significantly different across these regions (p= 0.43). The Funnel plot for assessing publication bias of *R*_*0*_ values computed from the random-effects model is shown in Fig.3. This Funnel plot is indicating the existence of asymmetry and publication bias. Additionally, the Egger’s test (P < 0.001) suggested the presence of small-study effects.

**Fig. 3.**
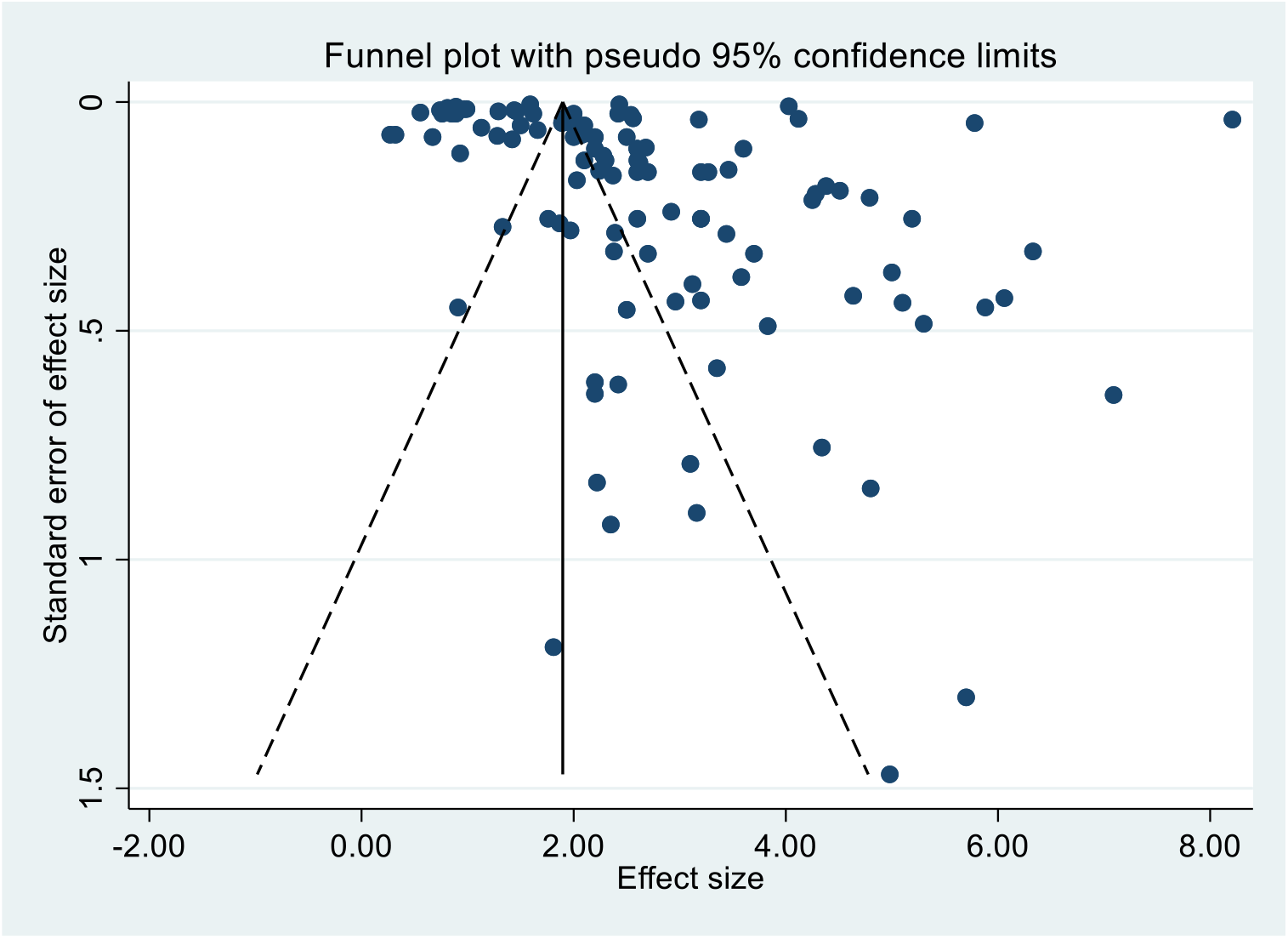
Funnel plot for the basic reproduction number (*R*_*0*_) values based on the random-effects model.

### Pooled estimation of the Case fatality rate (CFR)

The pooled estimation of the CFR from the random-effects model was 2.67 (2.25, 3.13). There was significant heterogeneity between studies (I^2^: 99.95, Q = 315324.53 with p = 0.00) (Table 2, and Fig. 4). The high value of *I*^*2*^ statistic (99.95%) confirms a high level of between-study heterogeneity.

**Table 2.** Effect size and test of heterogeneity for the case fatality rate (CFR) by region, population median age (median value), tests per thousand population (median value).

| Statistics | Region |  |  |  |  |  | Test for Subgroup differences |  |
| --- | --- | --- | --- | --- | --- | --- | --- | --- |
|  | Africa | Asia | Europe | North America | Oceania | South America |  |  |
| Effect Size (95% CI) | 7.11<br>(6.38, 7.91) | 2.51<br>(2.12, 2.95) | 2.66<br>(1.83, 3.64) | 3.40<br>(2.81, 4.04) | 3.45<br>(0.61, 17.18) | 2.89<br>(2.03, 3.89) | 131.42<br>(p-value = 0.00) | 2.67<br>(2.25, 3.13) |
| *I<sup>2</sup> statistic* | - | 99.66% | 99.98% | 99.84% | - | 99.69% |  | 99.95% |
| Cochran's Q statistics | - | - | - | - | - | - |  | 315324.53 |
| n (sample=158) | 1 | 72 | 60 | 15 | 1 | 9 |  | 158 |
| Total tests per thousand population |  |  |  |  |  |  |  |  |
|  | Less than or equal to 111.16 tests |  |  | Greater than 111.16 tests |  |  |  |  |
| Effect Size (95% CI) | 3.15 (2.57, 3.79) |  |  | 2.16 (1.57, 2.83) |  |  |  | 2.68 (2.25, 3.14) |
| *I<sup>2</sup> statistic* | 99.90% |  |  | 99.97% |  |  | 5.14<br>(p-value = 0.02) | 99.95% |
| Cochran's Q statistics | - |  |  | - |  |  |  | 315324.53 |
| n (sample=157) | 87 |  |  | 70 |  |  |  | 157 |
| Population Median Age Category |  |  |  |  |  |  |  |  |
|  | Less than or equal to 38.7 years |  |  | Greater than 38.7 years |  |  |  |  |
| Effect Size (95% CI) | 2.27 (1.85, 2.74) |  |  | 3.13 (2.44, 3.91) |  |  |  | 2.67 (2.25, 3.13) |
| *I<sup>2</sup> statistic* | 99.80% |  |  | 99.97% |  |  | 4.30<br>(p-value = 0.04) | 99.95% |
| Cochran's Q statistics | - |  |  | - |  |  |  | 315324.53 |
| n (sample=158) | 82 |  |  | 76 |  |  |  | 158 |

**Fig. 4.**
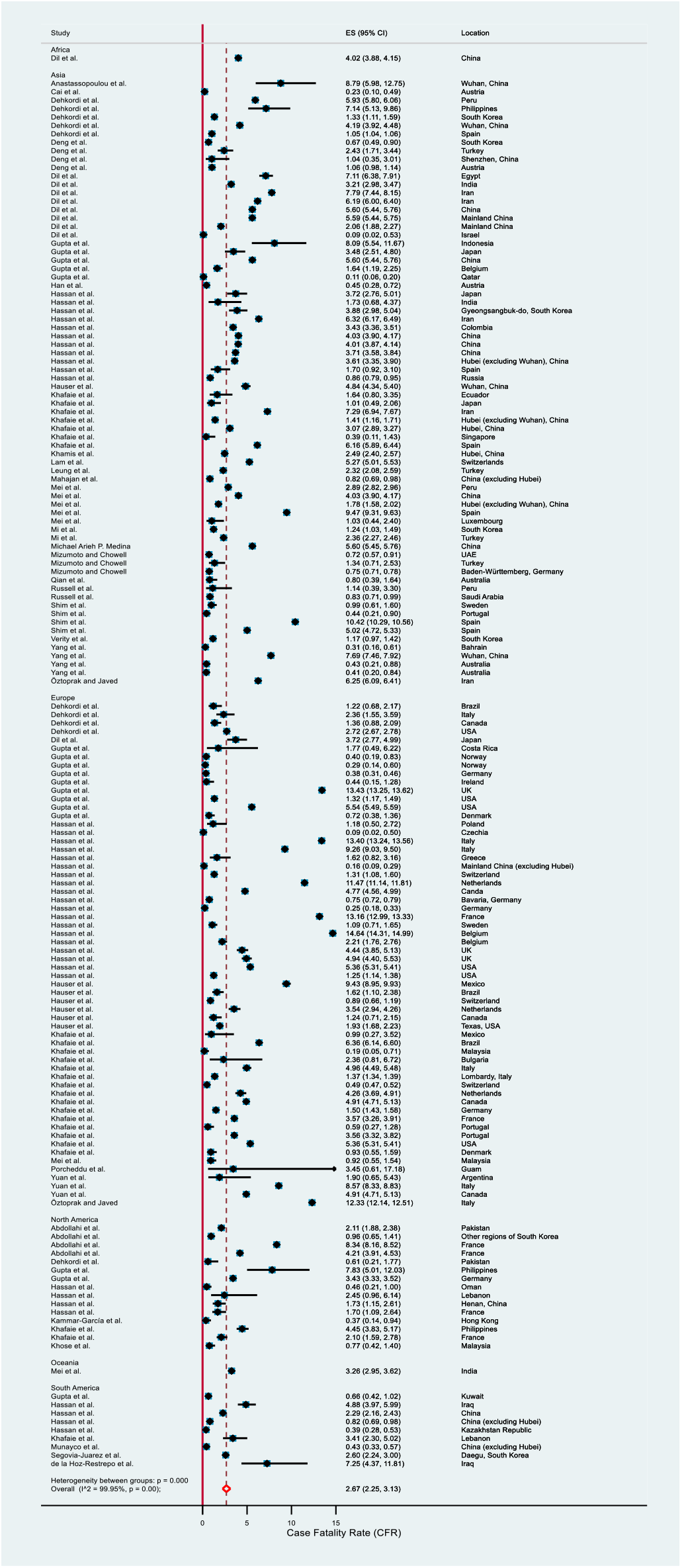
Forest plot of the case fatality rate (CFR) values based on the random-effects model.

The outcome of the subgroup analyses stratified by region showed that the CFR for Africa was 7.11 (6.38, 7.91), for Asia was 2.51 (2.12, 2.95), and for Europe was 2.66 (1.83, 3.64), for North America was 3.40 (2.81, 4.04), and for South America was 2.89 (2.03, 3.89) (Table 2). We observed higher mean CFR values for the countries with lower tests (3.15 vs. 2.16) and greater median population age (3.13 vs. 2.27). The subgroup difference tests using the random-effects model are significant (Table 2). This confirms that the estimated mean CFR values are significantly different across these subgroups. The Funnel plot for CFR values is shown in Fig. 5. Though the Funnel plot is indicating asymmetry, the Egger test (P= 0.182) suggested that there was no significant evidence of small-study effects.

**Fig. 5.**
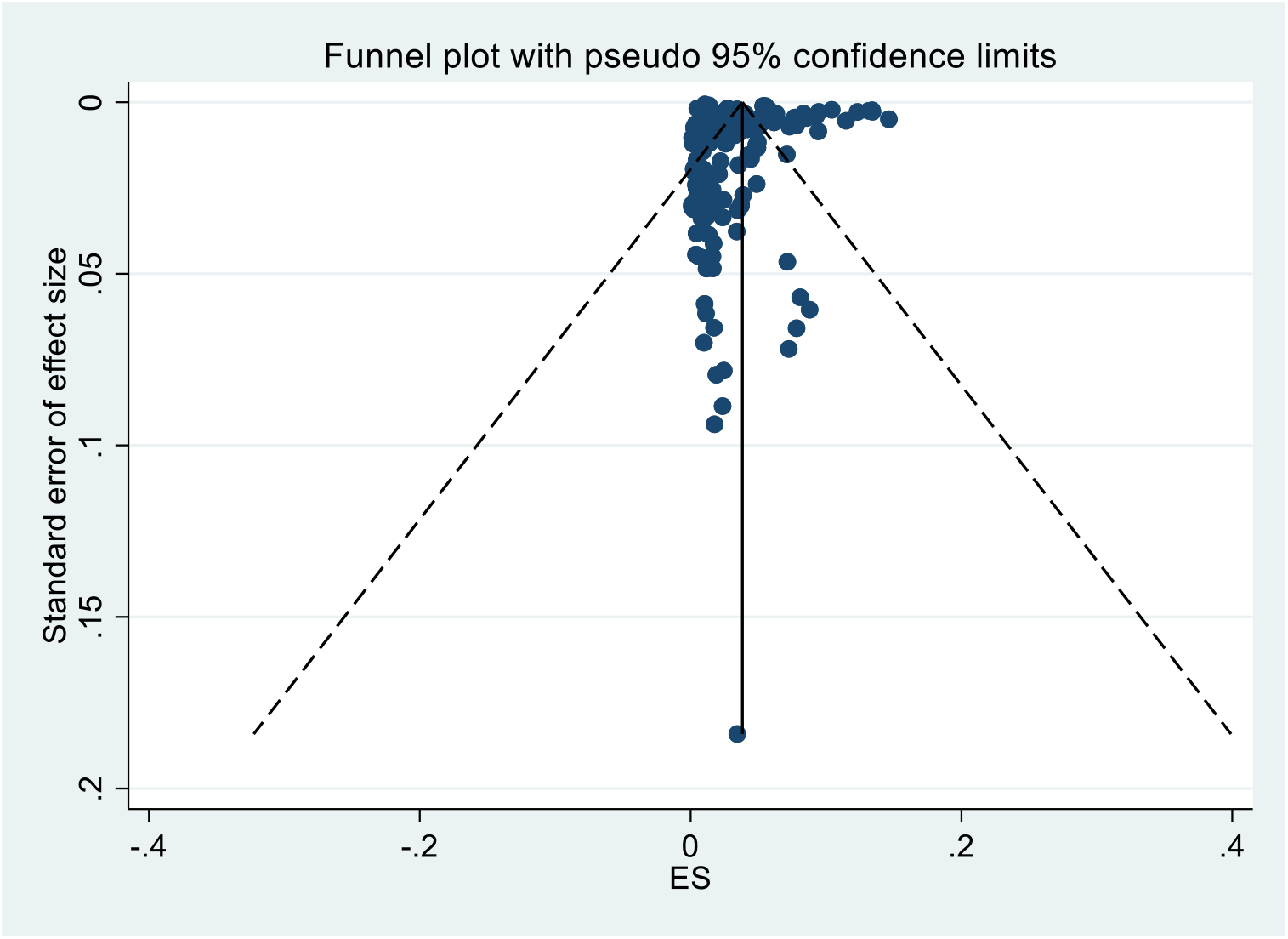
Funnel plot for the case fatality rate (CFR) values based on the random-effects model.

However, if we exclude the extreme CFR values (reported in the included articles), the pooled CFR estimation became 2.63 (2.24, 3.05). Furthermore, if we exclude the cases where naïve CFR (estimated as the ratio of the deaths to the confirmed cases mentioned in the article) > 4, the pooled CFR estimation became 1.23 (0.97, 1.53).

## Discussion

Our estimate of the *R*_*0*_ statistic implies that one infectious person can transmit the infection to more than two and a half susceptible persons in a naïve population in the absence of any control measures. However, the *R*_*0*_ did not vary significantly across the continents. Among these regions, the highest *R*_*0*_ was estimated for ‘North America’ (3.69) and lowest for ‘South America’ (2.32). We estimated a CFR of above two and a half, and the CFR varied in different continents (highest 7.11 in Africa, lowest 2.51 in Asia). We further observed lower CFR values of COVID-19 in the countries where a higher number of samples (per thousand population) were tested, and higher CFR values where the median age of the national population was higher (>38.7 years).

Our estimated overall *R*_*0*_ is slightly greater than WHO estimates of 1.4 to 2.5 [87] but smaller than the results of the other studies, i.e., He et al. and Alimohamadi et al. estimated *R*_*0*_ as 3.15 (95% CI: 2.41-3.90) and 3.32 (95% CI: 2.81 - 3.82), respectively [88,89]. Our estimation is similar to the *R*_0_ values estimated for the severe acute respiratory syndrome epidemic (*R*_0_= 2–3) but greater than the Middle East respiratory syndrome in the Middle East (*R*_*0*_ <1) [90]. However, high *R*_*0*_ values tend to be estimated in the early stages of epidemics and pandemics due to small sample sizes and incomplete information. Even if the overall *R*_*0*_ value in a population is low, the likelihood of transmission in some subgroups of that population may still be high as the total value of *R*_*0*_ in a population is the average of the *R*_*0*_ subtypes of that community.

Our estimated value for CFR is less than the WHO reported estimate of 3.4% [91] but similar to the estimates of the Chinese Center for Disease Control and Prevention (China CDC) (2.3%) [92] and He et al. (2.72% with 95% CI:1.29%-4.16%) [88]. Our estimated CFR is lower than the CFR values estimated for the other severe coronavirus, severe acute respiratory syndrome epidemic (9.6%), and the Middle East respiratory syndrome (40%) [90], but higher than that of influenza (0·1%) [93,94]. While multiple studies reported higher estimates of the case fatality rate than ours, those estimates are typically higher than the actual case fatality rate, as mild, atypical, and asymptomatic cases may not be identified (contrary to deaths), particularly in countries where access to testing is limited [95]. For example, before 11 March 2020 in the USA, sufficient, state-funded testing for all suspected cases was not available [96]. A study with 565 Japanese people who had been evacuated from Wuhan found that the number of infected people is likely to be about ten times greater than the number of registered cases. The CFR would then only be about one-tenth of that currently measured [97]. Our included studies may suffer from bias of these higher numbers of unreported cases, which could increase the pooled CFR estimation. Thus, we further estimated the CFR excluding the extreme values and the naive CFR greater than four. These gave us lower CFR values close to one.

In our estimation, the CFR varied among different regions, but the *R*_*0*_ did not vary significantly. We found lower CFR in the countries with a higher number of sample testing per thousand population. A higher number of testing allowed detection of more pre-/a-symptomatic and mild cases, which reduced the rate of fatal outcome [98]. We further found that countries with higher median age had higher CFR value. Older age has been recognized as one of the most common risk factors for a severe outcome of COVID-19 [99–101]. A study conducted with 5,484 patients in Italy found the CFR of COVID-19 was 0.43% for people below 70 years and 10.5% for people above 70 years and above [101]. Another study showed that an increase of 1% population aged 65 years and above increased the risk of death due to COVID-19 by 10% [100].

An estimate of *R*_*0*_ is affected by several factors, including the proportion of the susceptible population, population density, and awareness among people. Because SARS-COV-2 is a novel virus, we believe the proportion of the susceptible population remains the same across the world; however, other factors (density and awareness) vary. The density of the population is lower in European and North American countries compared with Asian counterparts. Thus, our estimation of the *R*_*0*_ for non-significant variation came as a surprise. However, the early phase of the pandemic produces incomplete data. Further studies on the *R*_*0*_ are essential to understand how different lockdown measures, especially face covering, social distancing, and demographic (population density) and environmental factors (temperature, humidity, UV light), affect the transmission dynamic of SARS-CoV-2 [102–105].

For developing and populated countries, the *R*_*0*_ value of 2.69 indicates the rapid spread of the COVID-19, and the CFR value of 2.67 is an indication of a relatively poor outcome of the treatments. Necessary precautions and strategies such as preventing mass gatherings, restricting the transportation system, and closing schools and universities should continue to prevent the outbreak of this disease until a successful vaccine arrived.

### Limitations

Some of the included studies were limited in terms of sample size, missing information on the onset of COVID-19, and data availability. Therefore, the reported findings should be interpreted cautiously within that context. Furthermore, our study was limited to the articles published in the English language only. Considering the epicenter of COVID-19, Chinese literature should be included in future systematic reviews. Another important limitation is the case detection and testing heterogeneity. Countries’ responses varied, and some countries are facing difficulties in testing enough samples. Further, the quality of the testing samples also varies in different countries. All these have affected the case detection and thus impacted the *R*_*0*_ and CFR. However, we believe the effects are minimum, and data across the world are still comparable.

## Conclusion

We estimated an *R*_*0*_ of 2.69 (95% CI: 2.40, 2.98) for SARS-CoV-2 and CFR of 2.67 (95% CI: 2.25, 3.13) for COVID-19 based on published literature from January to August 2020. The analysis by subgroups of regions confirmed a significant CFR variation, but the same was not found for the *R*_*0*_. Considering the estimated *R*_*0*_ and CFR values for COVID-19, implementation of the social distancing program and other non-pharmaceutical interventions are necessary steps to control the epidemic until an effective vaccine reaches to all susceptible population.

## Data Availability

The datasets generated and analyzed during this study are available from the corresponding author on reasonable request.

## Acknowledgement

We acknowledge the authors of the selected research articles. NH and RK is a member of the Pan-African Network on Emerging and Re-Emerging Infections (PANDORA-ID-NET – https://www.pandora-id.net/) funded by the European and Developing Countries Clinical Trials Partnership (EDCTP2) programme which is supported under Horizon 2020, the European Union’s Framework Programme for Research and Innovation. We also acknowledge Laura Macfarlane-Berry for reviewing and editing the manuscript and Md. Ahadur Rahman for his resource related support.

## Availability of data and materials

The full list of data and the data entries for all included studies is provided in the paper (Supplementary Tables S4 and S5). No additional supporting data is available.

## Ethical approval

Not required.

